# Accuracy of US CDC COVID-19 Forecasting Models

**DOI:** 10.1101/2022.04.20.22274097

**Authors:** Aviral Chharia, Govind Jeevan, Rajat Aayush Jha, Meng Liu, Jonathan M Berman, Christin Glorioso

**Author notes:** Authors claim equal contribution (Aviral Chharia, Govind Jeevan, Rajat Aayush Jha, Meng Liu).

## Abstract

Accurate predictive modeling of pandemics is essential for optimally distributing resources and setting policy. Dozens of case predictions models have been proposed but their accuracy over time and by model type remains unclear. In this study, we analyze all US CDC COVID-19 forecasting models, by first categorizing them and then calculating their mean absolute percent error, both wave-wise and on the complete timeline. We compare their estimates to government-reported case numbers, one another, as well as two baseline models wherein case counts remain static or follow a simple linear trend. The comparison reveals that more than one-third of models fail to outperform a simple static case baseline and two-thirds fail to outperform a simple linear trend forecast. A wave-by-wave comparison of models revealed that no overall modeling approach was superior to others, including ensemble models, and error in modeling has increased over time during the pandemic. This study raises concerns about hosting these models on official public platforms of health organizations including the US-CDC which risks giving them an official imprimatur and further raising concerns if utilized to formulate policy. By offering a universal evaluation method for pandemic forecasting models, we expect this work to serve as the starting point towards the development of more accurate models.

## Introduction

The COVID-19 pandemic (1) has resulted in at least 80.3 million confirmed cases and more than 1 million deaths in the United States alone. Worldwide, cases exceed 500 million, with at least 6 million deaths (2). The pandemic has affected every country and continues to present a major threat to global health. This has caused a critical need to study the transmission of emerging infectious diseases in order to make accurate case forecasts, especially during disease outbreaks. Case prediction models are useful for developing pandemic preventive and control methods, such as suggestions for healthcare infrastructure needs, isolation of infected persons, and contact activity tracking. Accurate models can allow better decision-making about the degree of precautions necessary for a given region at a particular time, which regions to avoid travel to and the degree of risk in various activities like public gatherings. Likewise, models can be used to proactively prepare for severe surges in cases by allocating resources such as oxygen or personnel. Collecting and presenting these models gives public health officials, and organizations such as the United States Centers for Disease Control and Prevention (CDC) (3), a mechanism to disseminate these predictions to the public, but risks giving them an official imprimatur, suggesting that these models were either developed by a government agency or endorsed by them.

Since the start of the pandemic, dozens of case prediction models for the US have been designed using a variety of methods. Each of these models depend on data available about cases, derived from a heterogeneous system of reporting, which can vary by county and suffer from regional and temporal delays. For example, some counties may collect data over several days and make it public at once, which creates an illusion of a sudden burst of cases. In counties with less robust testing programs, the lack of data can limit modeling accuracy. These methods are not uniform or standardized between groups that perform data collection, resulting in unpredictable errors. Underlying biases in the data, such as under-reporting, can produce predictable errors in model quality, requiring models to be adjusted to predict future erroneous reporting rather than actual case numbers. Such under-reporting has been identified by serology data (4; 5). Moreover, there is no universally agreed upon system for assessing and comparing the accuracy of case prediction models. Often published models use different methods, which makes direct comparisons difficult. The CDC has taken in data of case prediction models in a standardized way which makes direct comparisons possible (3). In this study, we use Mean Absolute Percent Error (MAPE) compared to the true case numbers to compare models for normalization purposes. First, we consider which models have the most accurately predicted case counts with the least MAPE. Next, we divide these models into five broad sub-types based on approach, i.e., epidemiological (or compartment) models, machine learning approaches, ensemble approaches, hybrid approaches, and other approaches, and compare the overall error of models using these approaches. We also consider which exclusion criteria might produce ensemble models with the greatest accuracy and predictive power.

A few studies (6; 7) have compared COVID-19 case forecasting models. However, the present study is unique in several aspects. First, it is focused on prediction models of US cases and takes into consideration all CDC models that pass the set inclusion criterion. Second, since these models were uploaded in a standardized format they can be compared across several dimensions such as *R*_0_, peak timing error, percent error, and model architecture. Third, we seek to answer several unaddressed questions relevant to pandemic case modeling. First, can we establish a metric to uniformly evaluate pandemic forecasting models? Second, What are the top-performing models during the four COVID-19 waves in the US and how do these fare on the complete timeline? Third, are there categories or classes of models that perform significantly better than others? Fourth, how do model predictions fare with increased forecast horizons? and lastly, how do the models compare to two simple baselines?

## Results

When case counts predicted by the various US CDC COVID-19 case prediction models are overlaid real world data, visually several features stand out as illustrated in Fig. 1a. On aggregate, models tend to approach the correct peak during various waves of the pandemic. However, some models undershot, others overshot, and many lagged the leading edge of real-world data by several weeks. The MAPE values of all US CDC models was analysed over the complete timeline and compared to two “Baselines”, which represented either an assumption that case counts would remain the same as the previous week (Baseline-I) or a simple linear model following the previous week’s case counts (Baseline-II) (see Fig. 1b). Here, ‘IQVIA_ACOE-STAN’, ‘USACE-ERDC_SEIR’, ‘MSRA-DeepST’, and ‘USC-SI_kJalpha_RF’ achieve the best performance with low MAPE ranging from 5% to 35%. In a comparison of overall performance, ensemble models performed significantly better than all other model types. However, the performance of ensemble models was not “significantly” better than the baseline models (no change or simple linear model) which performed better than both machine learning and epidemiological models overall (see Fig. 1c).

**Fig. 1.**
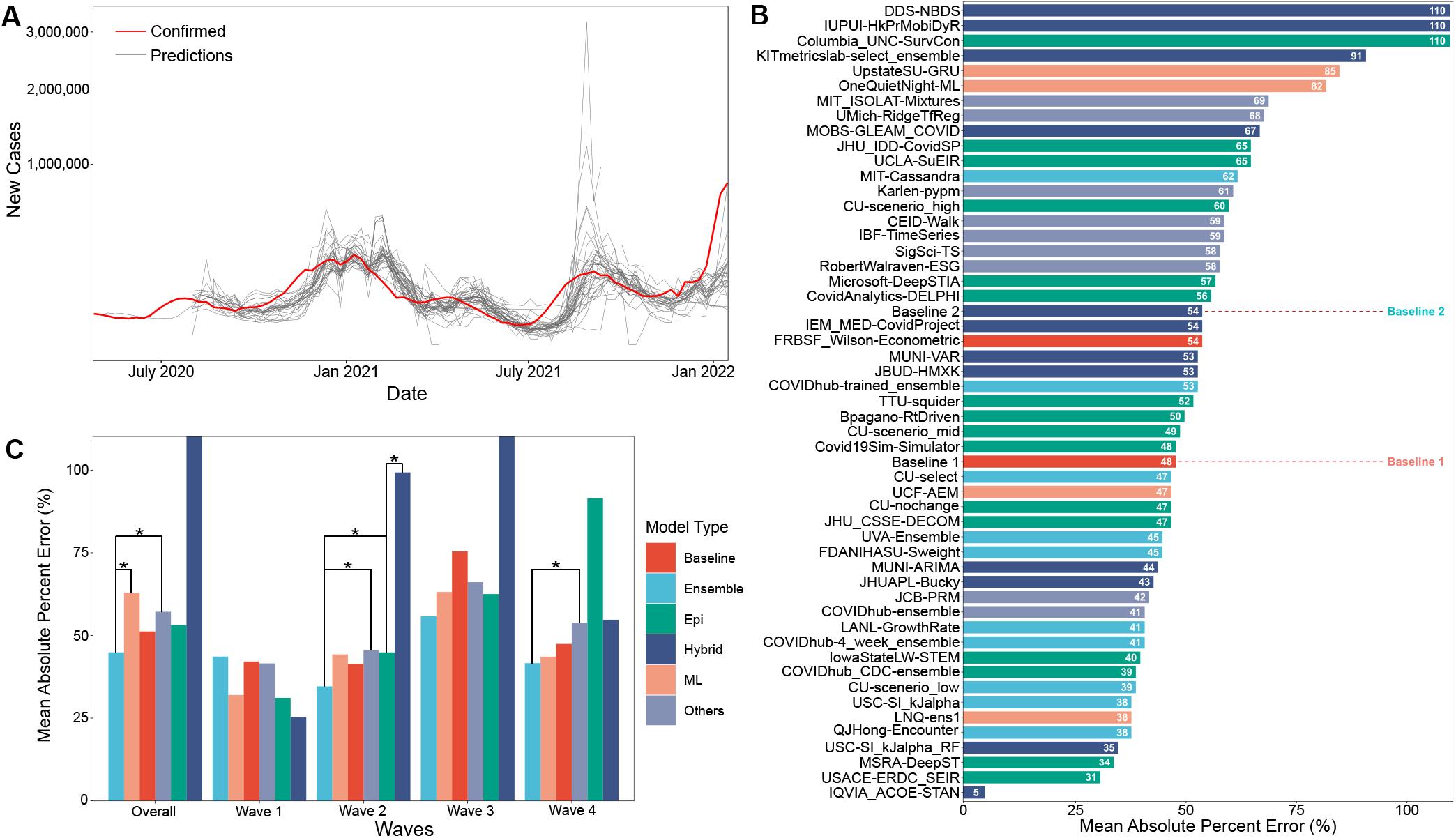
(A) Visual overlay of real case counts and predicted case counts across all waves examined. Actual case counts are shown in red, predicted counts are shown in grey with each trace representing a different CDC COVID-19 forecasting model on the sqrt plot. (B) MAPE values of US-CDC case prediction models on the complete timeline, i.e., Wave-I to IV. The y-axis is sorted descending from lowest error to highest. The color scheme represents the model category. (C) Bar graph showing the paired t-test results. Category-wise error achieved by the models on both overall and wave-wise from wave-1 to wave-4.

The MAPE values of all US CDC models were also analysed wave-wise (see Fig. 2). During the first wave of the pandemic, ‘Columbia_UNC-SurvCon’ achieved the lowest MAPE = 14%, closely followed by ‘USACE-ERDC_SEIR’ (MAPE = 17%) and ‘CovidAnalytics-DELPHI’ (MAPE = 25%). He, only 4 models performed better than both baselines. Three of these were epidemiological models while one was a hybrid model. From the t-test, on MAPE values of models categorized on the basis of model type (refer Fig. 1c), it can be inferred that during the first wave, hybrid models performed the best and attained the lowest MAPE. This was followed by the epidemiological models and those based on machine learning. On the other hand, ensemble models had the largest MAPE during this wave and none of them surpassed the Baseline-I MAPE, i.e., 31%. During the second wave, ‘IQVIA_ACOE-STAN’ performed the best with an MAPE score of 5% (see Fig. 2). In this wave, a total of 13 models performed better than both baselines, with MAPE ranging from 5 to 37. These included 5 ensemble models, 4 epidemiological models, 2 machine learning models and 2 hybrid models. All ensemble models exceed Baseline-I performance (that had MAPE = 37%), with the exception of ‘UVA-Ensemble’. The epidemiological models showed a staggered MAPE distribution. Followed by the hybrid and the models categorized as ‘other’ model sub-types, these have the lowest average MAPE in wave-II. In contrast to wave-I, ensemble models provide the best forecasts in wave-II (see Fig. 1c). Here, hybrid models are the worst performing models. During wave-III (see Fig. 1c), ensemble models performed similarly to wave-I. Baseline models had a relatively elevated high MAPE with Baseline-I and II MAPE scores being 74% and 77% respectively. In wave-III, ‘USC-SI_kJalpha’ is the best-performed model with MAPE= 32% (see Fig. 2). Here, 32 models performed better than both baseline models. These included 12 compartment models, 3 machine learning models, 4 hybrid models, 8 ensemble models, and 5 un-categorized models. In wave-IV of the pandemic, a number of models performed similarly between a MAPE of 28% and baseline of 47% (Fig. 2). Ensemble models performed the best whereas epidemiological models had the highest MAPE during this wave. Baseline-I and II MAPE scores were 47% and 48% respectively. In wave-IV, ‘LANL-GrowthRate’ is the best performed model with MAPE= 28% (Fig. 2). In the fourth wave, 17 models performed better than both baseline models. These included 6 compartment models, 7 ensemble models, 1 machine learning model, 2 hybrid models, and 1 uncategorized model.

**Fig. 2.**
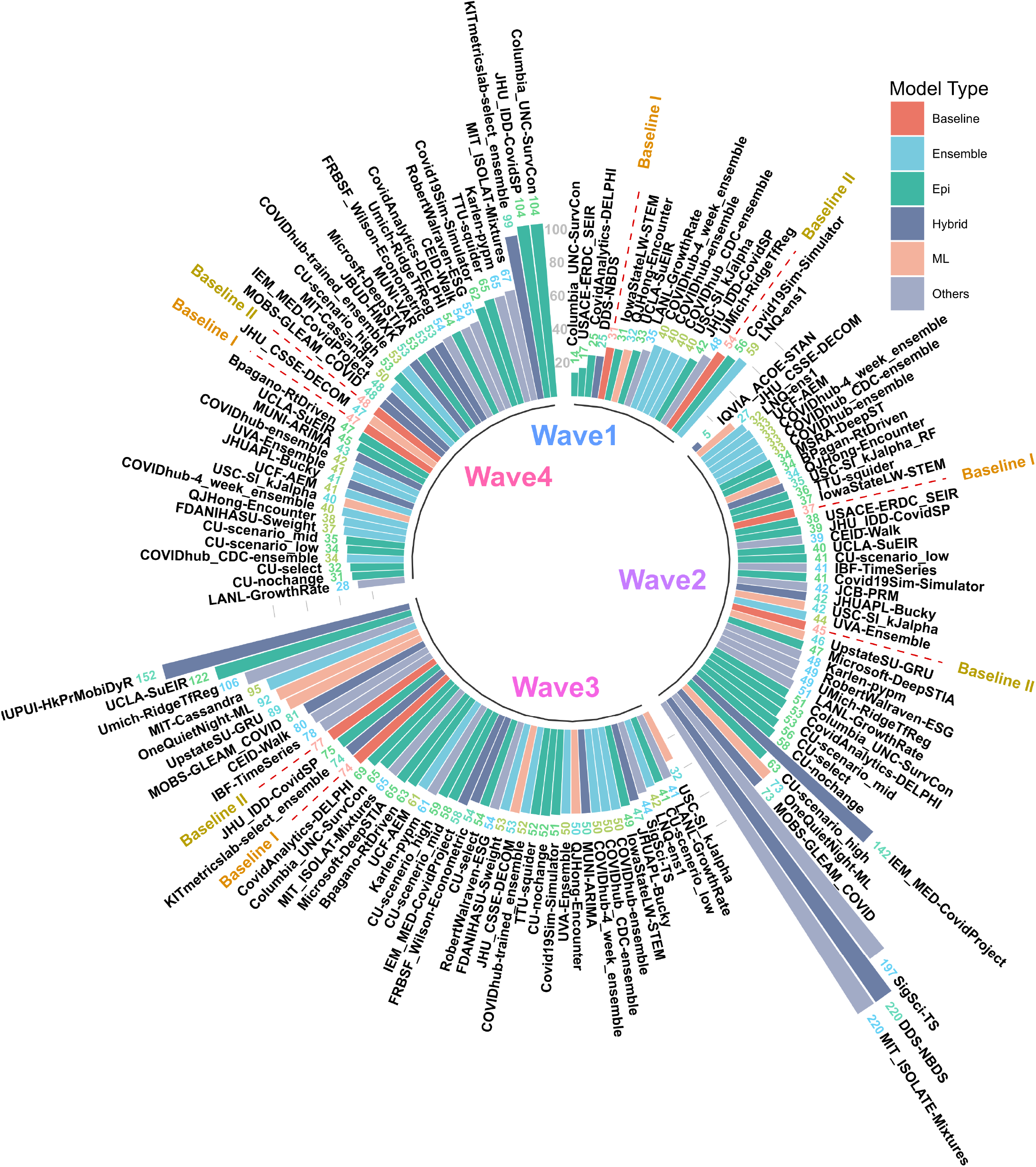
MAPE values of US CDC Case Prediction models in wave-I to IV. Models are sorted in descending order of MAPE. The color scheme represents the model category. Here “Baselines” are represented in red.

The MAPE of models in the US CDC database increased each week out from the time of prediction. Fig. 3 depicts a strictly increasing rise in MAPE with the increase in the forecast horizon. In other words, the accuracy of predictions declined the further out they were made. At one week from the time of prediction, the MAPE of models examined clustered just below 25% MAPE and declined to about 50% MAPE by four weeks. The MAPE in each week was relatively bi-modal, with several models fitting within a roughly normal distribution and others having a higher MAPE. The distribution of the points indicates that the majority of models project similar predictions for smaller forecast horizons, while the predictions for larger horizons are more spread out. The utility of model interpretation would improve by excluding those models that fall more than one standard deviation (*σ*) from the average MAPE of models.

**Fig. 3.**
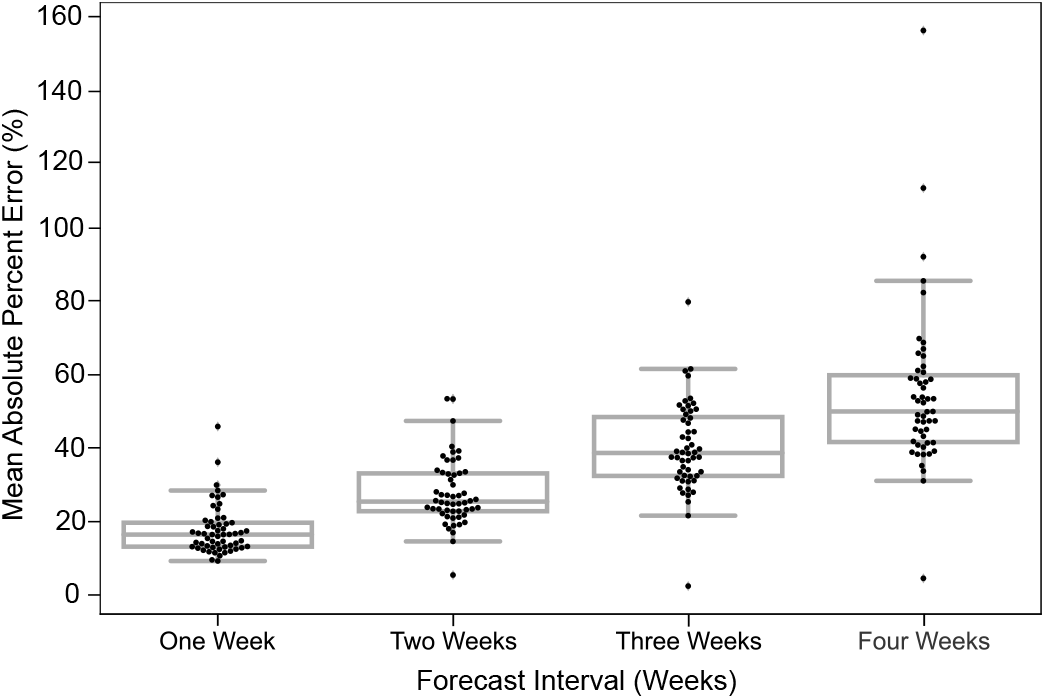
Predictions are most accurate closest to the time of prediction. The MAPE in predictions of all models for different forecast horizons is shown. The dots in each box plot represent the MAPE over all the predictions of a certain model for the corresponding forecast horizon. The y-axis is the MAE between the predicted case count and the reported case count. The x-axis is the forecast horizon.

Though we examined 54 models overall (reported 51 in the full timeline), many of these did not make predictions during the first wave of the pandemic when data was less available and therefore do not appear in the wave-wise MAPE calculation and subsequent analysis. The number of weeks for which each model provides predictions were highly variable, as seen in Fig. 4.

**Fig. 4.**
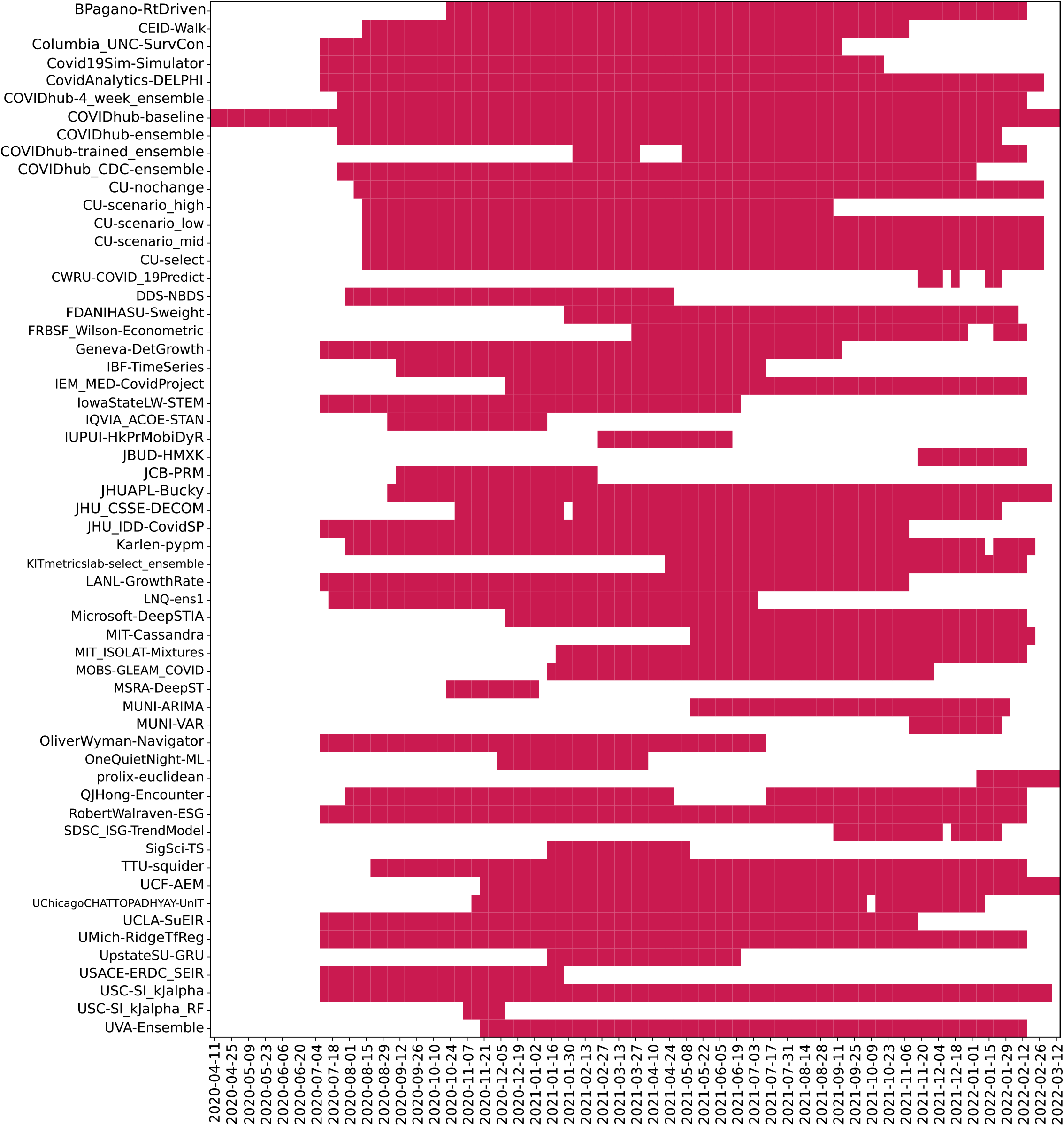
Bar plot depicting frequency of 04 week ahead predictions made by models. Here, models were ordered alphabetically on the y-axis. The x-axis represents target dates for which the predictions were made. Dates range from July 2020 to Jan 2022.

## Discussion

Accurate modeling is critical in pandemics for a variety of reasons. Policy decisions need to be made by political entities which must follow procedures, sometimes requiring weeks for a proposed policy intervention to become law and still longer to be implemented. Likewise, public health entities such as hospitals, nurseries, and health centers need “lead time” to distribute resources such as staffing, beds, ventilators, and oxygen supplies. However, modeling is limited, especially by the availability of data, particularly in early outbreaks (8). Resources such as ventilators are often distributed heterogeneously (9), leading to a risk of unnecessary mortality. Similarly, the complete homogeneous distribution of resources like masks is generally sub-optimal and may also result in deaths (10). Likewise, it is important to develop means of assessing which modeling tools are most effective and trustworthy. For example, a case forecasting model that consistently makes predictions that fare worse than assuming that case counts will remain unchanged or that they will follow a simple linear model isn’t likely to be useful in situations where modeling is critical. The use of these baselines allows for the exclusion of models that “fail” to predict case counts adequately.

Direct comparison of error based on the difference from real-world data potentially excludes important dimensions of model accuracy. For example, a model that accurately predicts the time course of disease cases (but underestimates cases by 20% at any given point) might have greater utility for making predictions about when precautions are necessary when compared to a model that predicts case numbers with only a 5% error but estimates peak cases two weeks late. To assess the degree of timing error, the peak of each model was compared to the true peak of cases that occurred within the model time window. MAPE i.e., the ratio of the error between the true case count and a model’s prediction to the true case count, is a straightforward way of representing the quality of predictive models and comparing between model types. This measure has some advantages over other methods of measuring forecast error. Because it deals with negative residuals by taking an absolute value rather than a squared value, reported errors are proportional. Percent error is also conceptually straightforward to understand. However, MAPE has advantages over absolute error. A model that predicts 201, 000 cases when 200, 000 cases occur has good accuracy but the same absolute error as a model that predicts 1100 cases when 100 occur. MAPE accounts for this by normalizing population size.

The average MAPE of successful models which we define as lower MAPE than either baseline model varied between methods depending on the wave they were measured in. In the first wave, epidemiological models had a mean MAPE of 31%, and machine learning models had a mean MAPE of 32%. In the second wave, these were 45% and 44% respectively. In the third wave, these were 63% and 63%, and in the fourth wave, these were 92% and 44% respectively. Therefore, we notice that the mean MAPE of models got worse with each wave. This is because each model type is susceptible to changing real-world conditions, such as the emergence of new variants with the potential to escape prior immunity, or a higher R_0_, new masking or lockdown mandates, the spread of conspiracy theories, or the development of vaccines which will decrease the number of individuals susceptible to infection.

In waves-2 and 3, the best performing model was a hybrid and machine learning model respectively. In waves 1 and 4, the best model was an epidemiological and an “other” model respectively (see Fig. 2). In each wave, some examples of each model type were successful, and some were unsuccessful. After wave 1, ensemble and ML models on average had the lowest MAPE but no model category significantly outperformed baseline models (Fig. 1c). These results suggest that no overall modeling technique is inherently superior for predicting future case counts.

Compartmental (or epidemiological) models broadly use several “compartments” which individuals can move between such as “susceptible,” “infectious,” or “recovered,” and use real-world data to arrive at estimates for the transition rate between these compartments. However, the accuracy of a compartment model depends heavily on accurate estimates of the R_0_ in a population, a variable that changes over time, especially as new virus variants emerge. For example, the emergence of the Iota variant of SARS-CoV-2 (also known as lineage B.1.526) resulted in an unpredicted increase in case counts (11). Machine learning models train algorithms that would be difficult to develop by conventional means. These models “train” on real-world data sets and then make predictions based on that past data. Machine learning models are sensitive to the datasets they are trained on, and small or incomplete datasets produce unexpected results. Hybrid models make use of both compartment modeling and machine learning tools. On the other hand, ensemble models combine the results of multiple other models hoping that whatever errors exist in the other models will “average out” of the combined model. Ensemble models potentially offer an advantage over individual models in that by averaging the predictions of multiple models, flawed assumptions or errors in individual models may “average out” and result in a more accurate model. However, if multiple models share flawed assumptions or data, then averaging these models may simply compound these errors. An individual model may achieve a lower error than multiple flawed models in these cases.

The “baseline” models had a MAPE of 48% and 54% over the entire course of the pandemic in the US. Most models did not perform better than these. Among those that did, we discuss the five with the best performance (in increasing order of their performance). First, “QJHong-Encounter” is a model by Qijung Hong, an Assistant professor at Arizona State University. This model uses an estimate of encounter density (how many potentially infectious encounters people are likely to have in a day) to predict changes in estimated R (reproduction number), and then uses that to estimate future daily new cases. The model uses machine learning. It had a MAPE of 38% over all waves. Second, “USC-SI_kJalpha_RF” is a hybrid model from the University of Southern California Data Science Lab. This model also uses a kind of hybrid approach, where additional parameters are modeled regionally for how different regions have reduced encounters, and machine learning is used to estimate parameters (12). It had a MAPE of 35% over all waves. Third, “MSRA-DeepST” is a SIR hybrid model from Microsoft Research Lab-Asia that combines elements of SEIR models and machine learning. It had a MAPE of 34%. Next, “USACE-ERDC_SEIR” is a compartment model developed by the US Army Engineer Research and Development Center COVID-19 Modeling and Analysis Team. It adds to the classic SEIR model, adding additional compartments for unreported infections and isolated individuals. It used Bayesian estimates of prior probability based on subject matter experts to select initial parameters. It had a MAPE of 31%. Lastly, “IQVIA_ACOE_STAN” is a machine learning model from IQVIA-Analytics Center of Excellence and has the highest apparent performance on the overall timeframe. The calculated MAPE for this model was 5%. Notably, the MAPE of 5% is much lower than the MAPE of 31% of the next closest model.

Although MAPE is superior to other methods of comparing models, there are still some challenges. For example, “IQVIA_ACOE_STAN” appears to be the lowest model by far, but the only data available covers a relatively short time frame, and unfortunately discontinued its contribution to the CDC website after Wave 2. The time frame that it predicted also does not include any changes in case direction from upswings and downswings. This advantages the model compared to other models, which might cover time periods where case numbers peak or new variants emerge. This highlights a potential pitfall of examining this data: the models are not studying a uniform time window.

Data reporting is one of the sources of error affecting model accuracy. There is significant heterogeneity in reporting of COVID-19 cases by state. This is caused by varying state laws, resources made available for testing, the degree of sequencing being done in each state, and other factors. Additionally, different states have heterogeneity in vaccination rate, population density, implementation of masking and lockdown, and other measures which may affect case count predictions. Therefore, the assumptions underlying different models and the degree to which this heterogeneity is taken into account may result in models having heterogeneous predictive power in different states. Although this same thinking could be extended to the county level, the case count reporting in each county is even more variable and makes comparisons difficult.

To make the comparison between models more even, we used multiple times segments to represent the various waves during the pandemic. Models that have made fewer predictions, particularly avoiding the “regions of interest” such as a fresh wave or a peak, would only be subjected to a less challenging evaluation than the models that covered most of the timeline. Further, the utility of models with only a few predictions within “regions of interest” is also questionable. Considering these aspects, we define 4 time segments corresponding to each of the major waves in the US. Within each of these waves of interest, we consider only models that made a significant number of predictions for the purpose of comparison. Such a compartmentalized comparison is now straightforward, as all models within a time segment can now have a common evaluation metric.

We focus on evaluating the performance of models for their 4-week ahead forecasts. A larger forecast horizon provides a higher real-world utility in terms of policy-making or taking precautionary steps. We believe this to be a more accurate representation of the predictive abilities of models as opposed to smaller windows in which desirable results could be achieved by simply extrapolating the present trend. Therefore, due to the time delays associated with policy decisions and the movement of critical resources and people, the long-term accuracy of models is of critical importance. To take an extreme example, a forecasting model that only predicted one day in advance would have less utility than one that predicted ten days in advance.

The failure of roughly one-third of models in the CDC database to produce results superior to a simple linear model should raise concerns about hosting these models in a public venue. Without strict exclusion criteria, the public may not be aware that the are significant differences in the overall quality of these models. Each model type is subject to inherent weaknesses of the available data. The accuracy of compartment models is heavily dependent on the quality and quantity of reported data and also depends on a variable that might change with the emergence of new variants. Heterogeneous reporting of case counts, variable accuracy between states, and variable early access to testing resulted in limited data sets. Likewise, it seems that since training sets did not exist, machine learning models were unable to predict the Delta variant surge. Robust evidence-based exclusion criteria and performance-based weighting have the potential to improve the overall utility of future model aggregates and ensemble models.

Because the US CDC has a primary mission focused on the United States, the models included are focused on United States case counts. However, globally the assumptions necessary to produce an accurate model might differ due to differences in population density, vaccine availability, and even cultural beliefs about health. However, identifying the modeling approaches that work best in the United States provides a strong starting point for global modeling. Some of the differences in modeling will be accounted for by different input data, which can be customized by country or different training sets in the case of machine learning models.

The ultimate measure of forecasting model quality is whether the model makes a prediction that is used fruitfully to make a real-world decision. Staffing decisions for hospitals can require a lead time of 2-4 weeks to prevent over-reliance on temporary workers, or shortages (13). Oxygen has become a scarce resource during the COVID-19 pandemic, and also needs lead time (14). This has had real-world policy consequences as public officials have ordered oxygen imports well after there were needed to prevent shortages (15). Indeed for sufficient time to be available for public officials to enact new policies and for resources to be moved, a time frame of eight weeks is preferable. The need for accurate predictions weeks in advance is confounded by the declining accuracy of models multiple weeks in advance, especially considering the rise time of new variant waves (Fig. 3). During the most recent wave of infections, news media reported on the potential for the “Omicron” variant of SARS-CoV-2 to rapidly spread in November of 2021, however in the United States, an exponential rise did was not apparent in case counts until December 14th, when daily new case counts approximated 100k new cases per day, and by January 14th, 2022, new cases exceed 850, 000 new cases per day. The time when accurate predictive models are most useful is ahead of rapid rises in cases, something none of the models examined were able to predict, given the rise in SARS-CoV-2 cases that occurred during the pandemic.

Although forecasting models have gained immense attention recently, many challenges are faced in developing these to serve the needs of governments and organizations. We found that most models are not better than CDC baselines. The benchmark for resource allocation ahead of a wave still remains the identification and interpretation of new variants and strains by biologists. We propose a reassessment of the role of forecasting models in pandemic modeling. These models as currently implemented can be used to predict the peak and decline of waves that have already been initiated and can provide value to decision-makers looking to allocate resources during an outbreak. Prediction of outbreaks beforehand however still requires “hands-on” identification of cases, sequencing, and data gathering. The lack of clean, structured, and accurate datasets also affects the performance of the case prediction models, making the estimation of patient mortality count and transmission rate significantly harder. More robust sources of data on true case numbers, variants, and immunity would be useful to create more accurate models that the public and policy makers can use to make decisions.

## Methods

This work compares various US CDC COVID-19 forecasting models by their quantitative aspects evaluating their performance in strictly numerical terms over various time segments. The US CDC collects weekly forecasts for COVID cases in four different horizons: 1-week, 2-weeks, 3-weeks and 4-weeks, i.e., each week, the models make a forecast for new COVID cases in each of the four consecutive weeks from the date of the forecast. The forecast horizon is the length of time into the future for which forecasts are to be prepared. In the present study, we focus on evaluating the performance of models for their 4-week ahead forecasts.

### Datasets

The data for the confirmed case counts are taken from the COVID-19 Data Repository (https://github.com/CSSEGISandData/COVID-19/tree/master/csse_covid_19_data/csse_covid_19_daily_reports_us), maintained by the Center for Systems Science and Engineering at the Johns Hopkins University. The data for the predicted case counts, of all the models, is obtained from the data repository for the COVID-19 Forecast Hub (https://github.com/reichlab/covid19-forecast-hub), which is also the data source for the official US-CDC COVID-19 forecasting page. Both these datasets were preprocessed to remove unwanted data items. For plotting Fig. 1a, the *Pyplot* module from the *Matplotlib* library in Python was used.

### Categorizing Models

The models were categorized into five different categories-Ensemble, Epidemiological, Hybrid, and Machine Learning. This was based on keywords found in model names and going to each model description on their respective web-pages and articles. The models which did not broadly fall into these categories were kept in ‘others’. These models use very different methods to arrive at predictions. We are comprehensively looking at 51 models. The CDC also uses an ensemble model, and we looked at whether this was better than any individual model. For each model uploaded to the CDC website, MAPE was calculated and reported in this study, and the models were compared wave-wise as shown in various figures. For each model, the model type was noted, as well as the month proposed.

### Wave Definition

A thorough search reveals M.D., epidemiologists, and policymakers do share the same underlying principles of the term ‘wave’. Popular media explain “the word ‘wave’ implies a natural pattern of peaks and valleys”. WHO stated in order to say one wave is ended, the virus has to be brought under control and cases have to fall substantially, then for a second wave to start, you need a sustained rise in infections. As part of their National Forecasts for COVID cases, CDC has reported the results from a total of 54 different models at various instances of time during the pandemic. We define the waves, i.e., Wave-I: July, 6th 2020 to August 31st, 2020, Wave-II: September, 1st 2020 to February, 14th 2021, Wave-III: February, 15th 2021 to July, 26th 2021 and Wave-IV: July, 27th 2021 to January, 17th 2022, corresponding to each of the major waves in the US.

### Baselines

The performance of the models was evaluated against two simple baselines. Baseline-I is the ‘CovidHub-Baseline’ (or CDC’s baseline), i.e., the median prediction at all future horizons is the most recent observed incidence. Baseline-II is the linear predictor extrapolation using slope of change in reported active cases between the two weeks preceding date of forecast. These baselines are included in the bar charts (shown in Fig. 2 and Fig. 1b). Within each of the waves of interest, only models that made a significant number of predictions are considered for the purpose of comparison. We only consider models that have made predictions for at least 25% of the target dates covered by the respective time segment for all comparisons in this section. The MAPE was calculated on the four-weeks forecast horizon. Fig. 1b illustrates the performance of models across all the waves. Same procedure, as in Fig. 2, was followed for plotting Fig. 1b.

### Model Comparison

The performance of all the models are compared, wave-wise and on the complete timeline based on the MAPE (or mean absolute percent error). MAPE is defined as the ratio of absolute percentage errors of the predictions. Error refers to the difference between the confirmed case counts and the predicted case counts. Here, *n* = number of datapoints, *A*_*t*_ is the actual value and *P*_*t*_ denoted the predicted value.

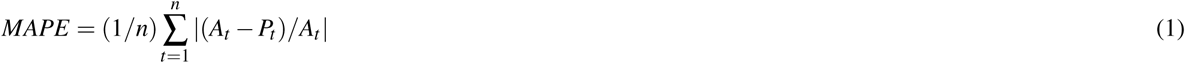

Fig. 1c shows the paired t-test results on the category-wise errors, achieved by the models on overall as well as wave-wise. The mean of the MAPE values is calculated for each category in model-type, for overall and each wave separately. The mean is calculated by adding the MAPE values of all the models in a category and dividing it by the number of models in that category for the corresponding wave. Then, pairwise t-test is performed to determine if there is a significant difference between the means of two groups. We used the *ttest_ind* function of the *statsmodels* module in Python to perform the test.

### Statistical Analysis

The statistical significance in all figure was determined by a two-tailed Students t-test p-value. Bar pots having p-value less than 0.05 (statistically significant), are joined using an asterisk.

In Fig. 3, box-plots are made representing the MAPE over all the predictions of a certain model for the corresponding forecast horizon. A box-plot displays the distribution of data based on a five number summary (“minimum”, first quartile (Q1), median, third quartile (Q3), and “maximum”). We used the boxplot function (of *seaborn* library) in python to plot it. *Seaborn* is a Python data visualization library based on *Matplotlib*.

## Data Availability

The data used in this study is publicly available from the COVID-19 Data Repository, maintained by the Center for Systems Science and Engineering (CSSE) at Johns Hopkins University. The data for the predicted case counts, of all the models, is obtained from the data repository for the COVID-19 Forecast Hub, the data source for the official US-CDC COVID-19 forecasting page. The source code that supports the findings of this research is available from the corresponding author upon request.

https://github.com/CSSEGISandData/COVID-19/tree/master/csse_covid_19_data/csse_covid_19_daily_reports_us

https://github.com/reichlab/covid19-forecast-hub

## Author contributions statement

A.C. worked on literature survey, wrote first draft and managed the project. G.J. worked on the code for pulling CDC-model predictions data and calculating MAPE. R.J. and M.L. prepared the manuscript figures. J.B. improved the discussion section, provided feedback and contributed to the development of the manuscript. C.G. thought of the idea of the project, supervised the project, helped to draft, review and edit the manuscript. All authors have read the final manuscript.

## Data Availability

The data used in this study is publicly available from the COVID-19 Data Repository, maintained by the Center for Systems Science and Engineering at Johns Hopkins University (https://github.com/CSSEGISandData/COVID-19/tree/master/csse_covid_19_data/csse_cov The data for the predicted case counts, of all the models, is obtained from the data repository for the COVID-19 Forecast Hub (https://github.com/reichlab/covid19-forecast-hub), the data source for the official US-CDC COVID-19 forecasting page.

## Code Availability

The source code that supports the findings of this research is available from the corresponding author upon request.

## Extended Data

The extended data section attached with the manuscript contains additional figures and tables that support the main content.

**Fig. S.1.**
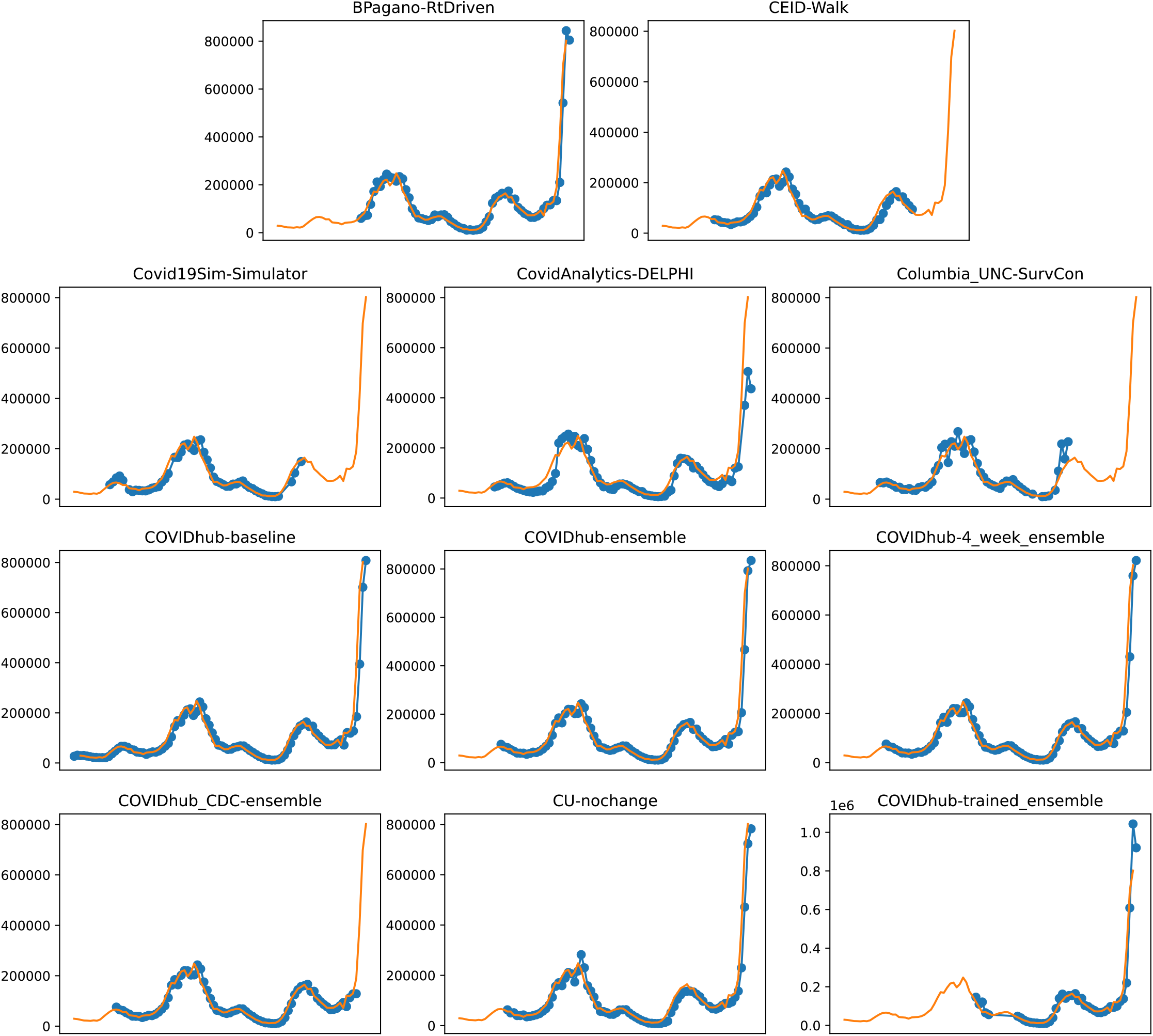
Plots depicting the peak predictions of US-CDC models-‘BPagano-RtDriven’, ‘CEID-Walk’, ‘Covid19Sim-Simulator’, ‘CovidAnalytics-DELPHI’, ‘Columbia_UNC-SurvCon’, ‘COVIDhub-baseline’, ‘COVIDhub-ensemble’, ‘COVIDhub-4_week_ensemble’, ‘COVIDhub_CDC-ensemble’, ‘CU-nochange’, ‘COVIDhub-trained_ensemble’, over complete timeline

**Fig. S.2.**
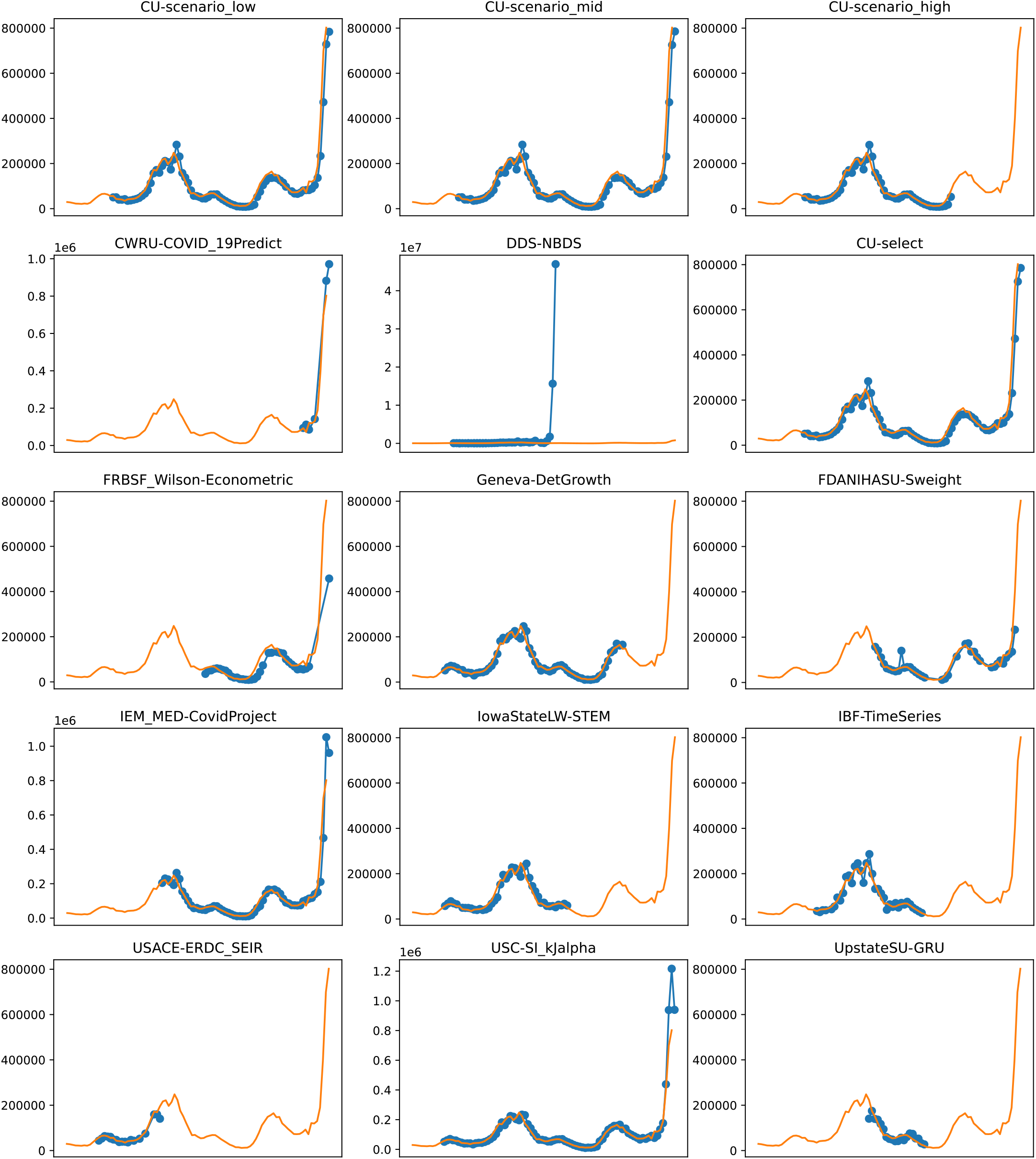
Plots depicting the peak predictions of various US-CDC models - ‘CU-scenerio_low’, ‘CU-scenario_mid’, ‘CU-scenerio_high’, ‘CWRU-COVID_19Predict’, ‘DDS-NBDS’, ‘CU-select’, ‘FRBSF_Wilson-Econometric’, ‘Geneva-DetGrowth’, ‘FDANIHASU-Sweight’, ‘IEM_MED-CovidProject’, ‘IowaStateLW-STEM’, ‘IBF-TimeSeries’, ‘USACE-ERDC_SEIR’, ‘USC-SI_KJalpha’, ‘UpstateSU-GRU’, over complete timeline

**Fig. S.3.**
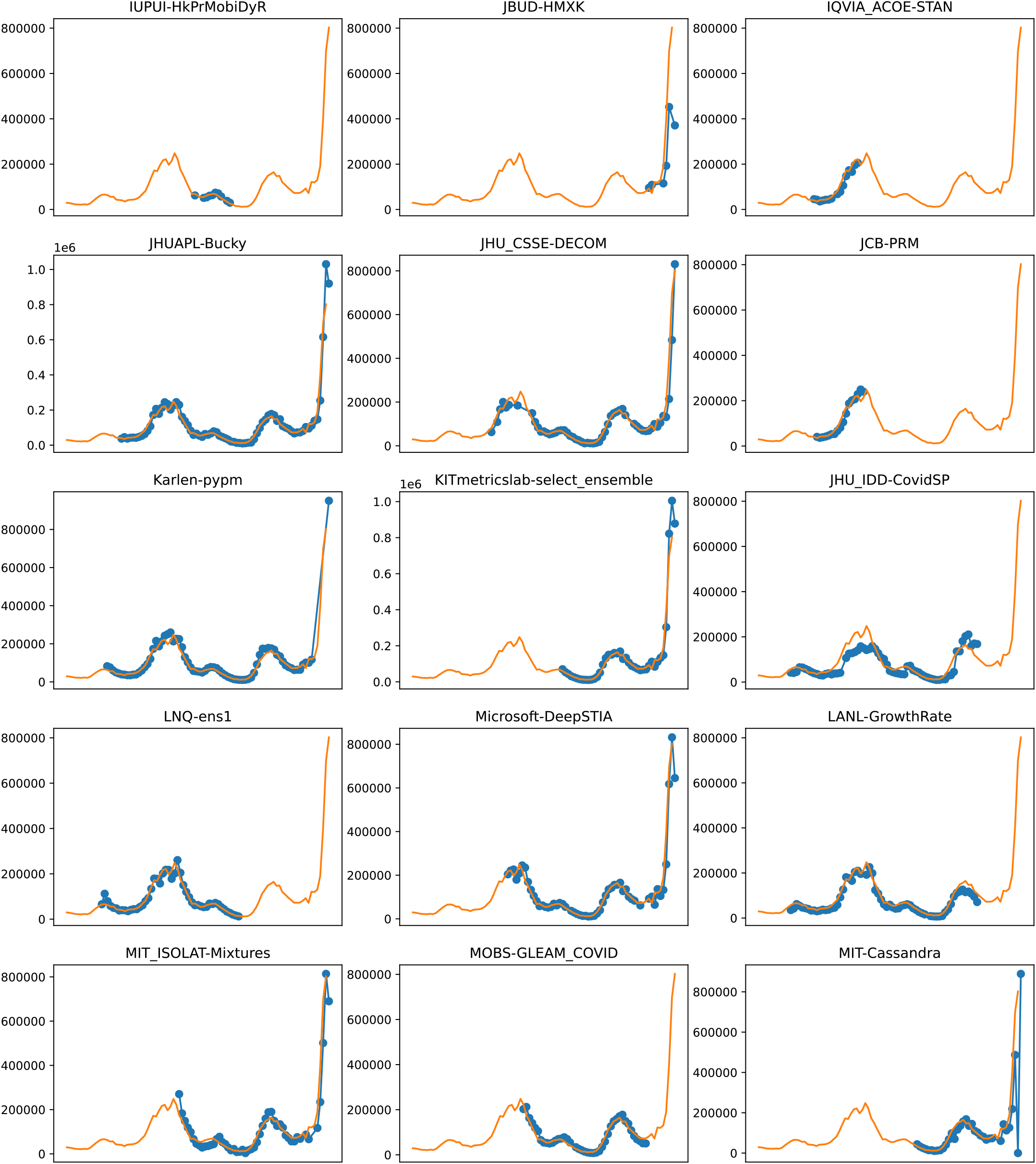
Plots depicting the peak predictions of various US-CDC models - ‘IUPUI-HkPrMobiDyR’, ‘JBUD-HMXK’, ‘IQVIA_ACOE-STAN’, ‘JHUAPL-Bucky’, ‘JHU_CSSE-DECOM’, ‘JCB-PRM’, ‘Karlen-pypm’, ‘KITmetricslab-select_ensemble’, ‘JHU_IDD-CovidSP’, ‘LNQ-ens1’, ‘Microsoft-DeepSTIA’, ‘LANL-GrowthRate’,’MIT_ISOLAT-Mixtures’, ‘MOBS-GLEAM_COVID’, ‘MIT-Cassandra’, over complete timeline

**Fig. S.4.**
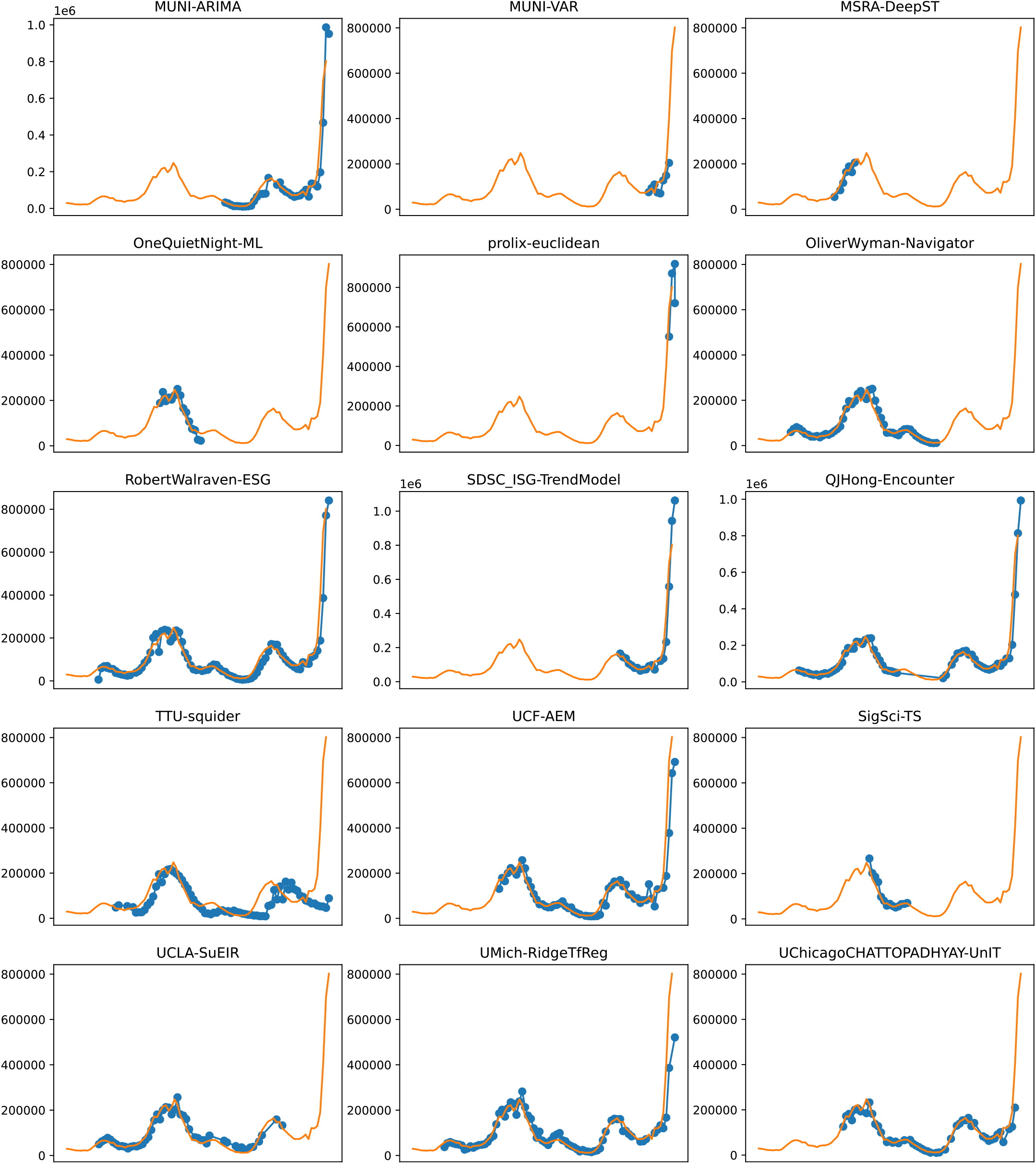
Plots depicting the peak predictions of various US-CDC models - ‘MUNI-ARIMA’, ‘MUNI-VAR’, ‘MSRA-DeepST’, ‘OneQuietNight-ML’, ‘prolix-euclidean’, ‘OliverWyman-Navigator’, ‘RobertWalraven-ESG’, ‘SDSC_ISG-TrendModel’, ‘QJHong-Encounter’, ‘TTU-squider’, ‘UCF-AEM’, ‘SigSci-TS’, ‘UCLA-SuEIR’, ‘UMich-RidgeTfReg’ and ‘UChicagoCHATTOPADHYAY-UnIT’ over complete timeline

**Table S1.**
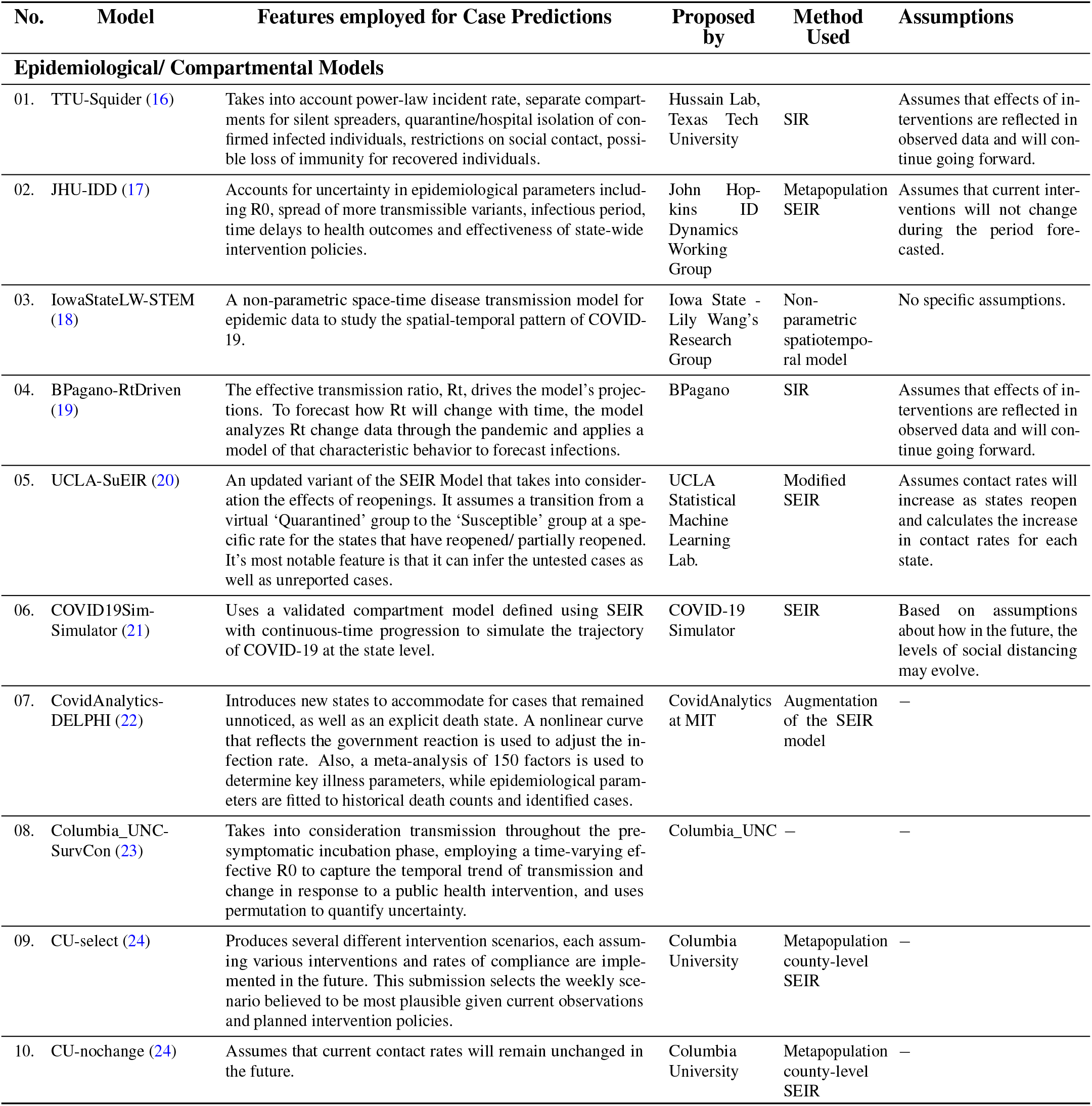
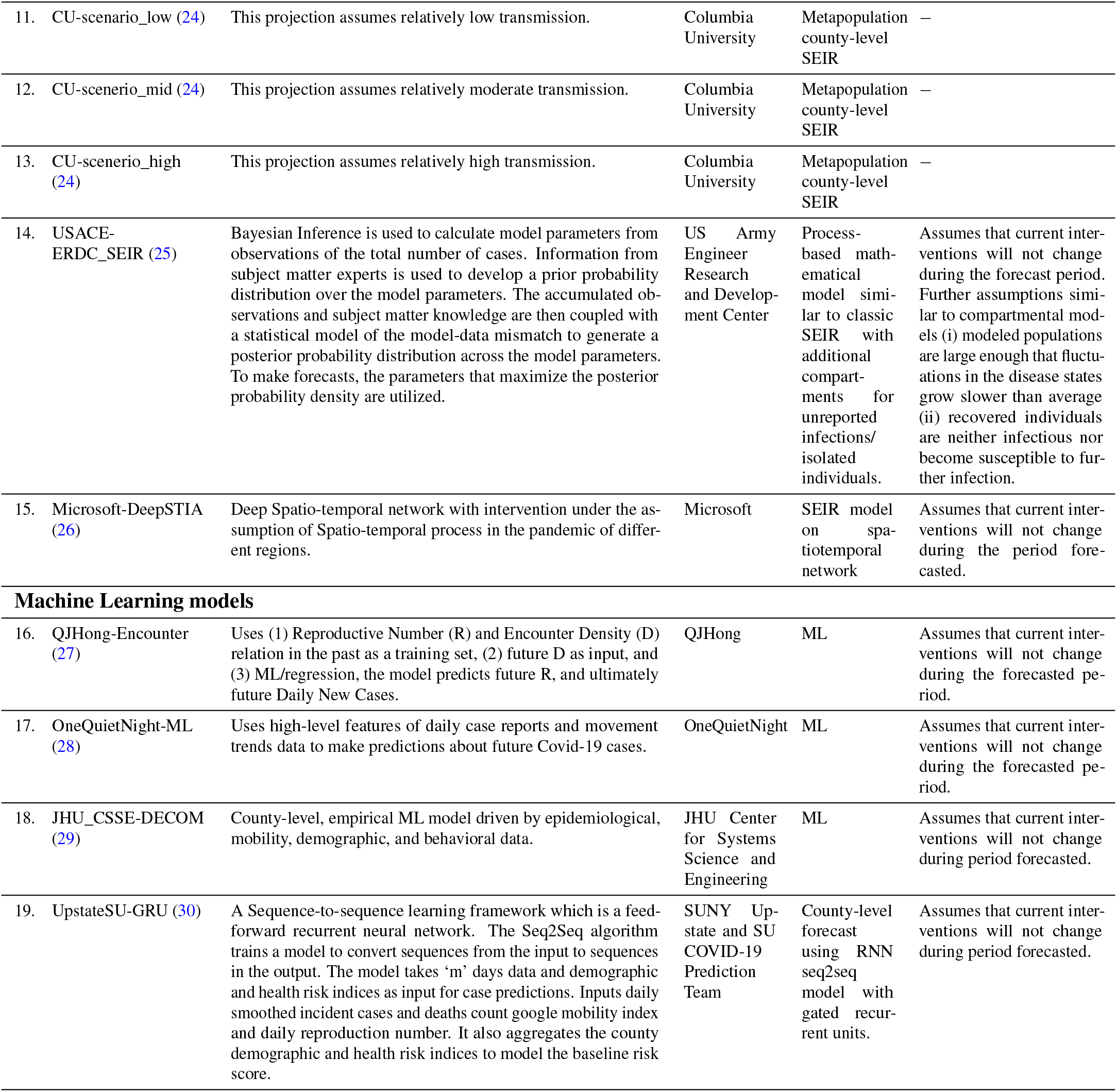
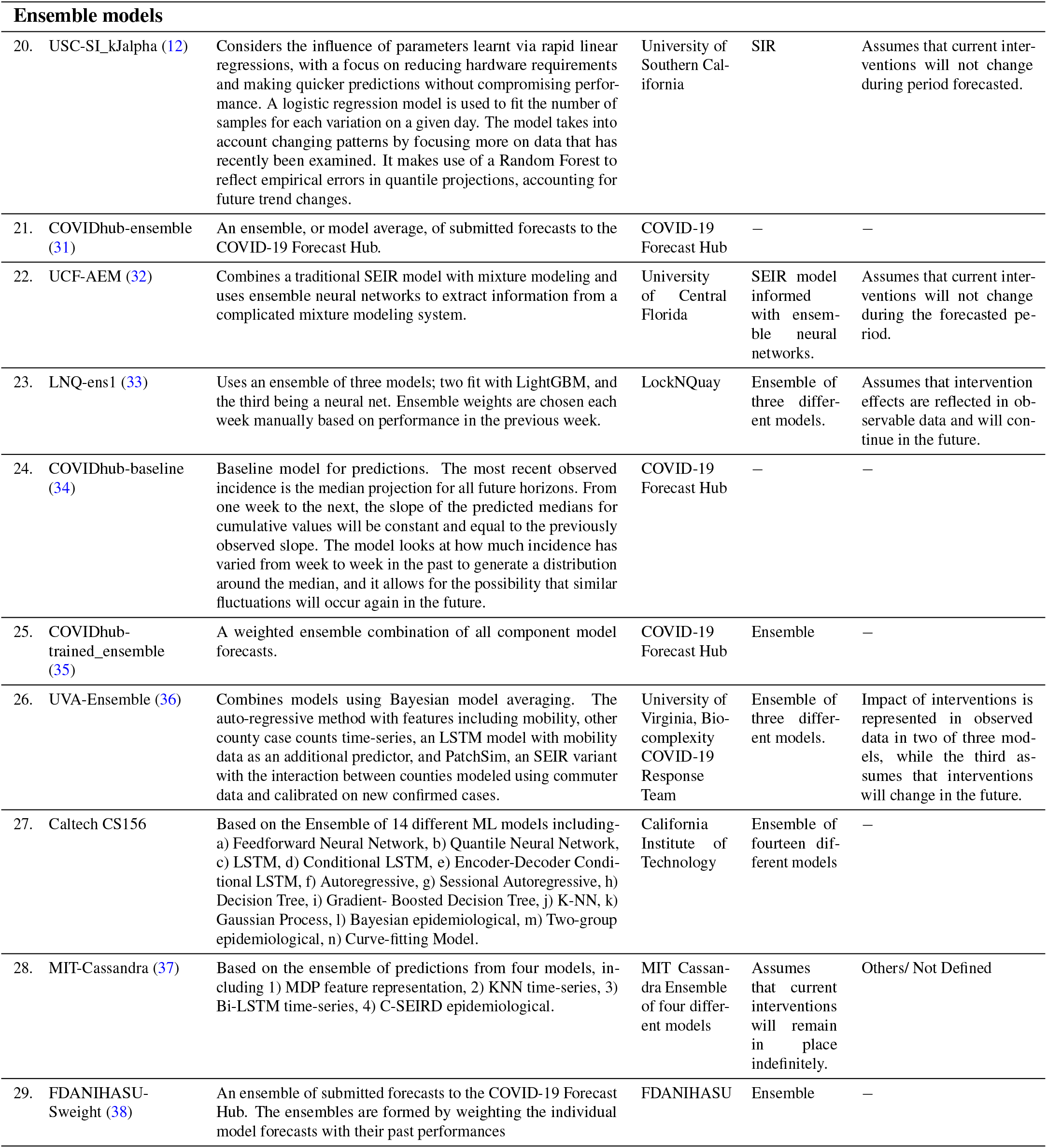
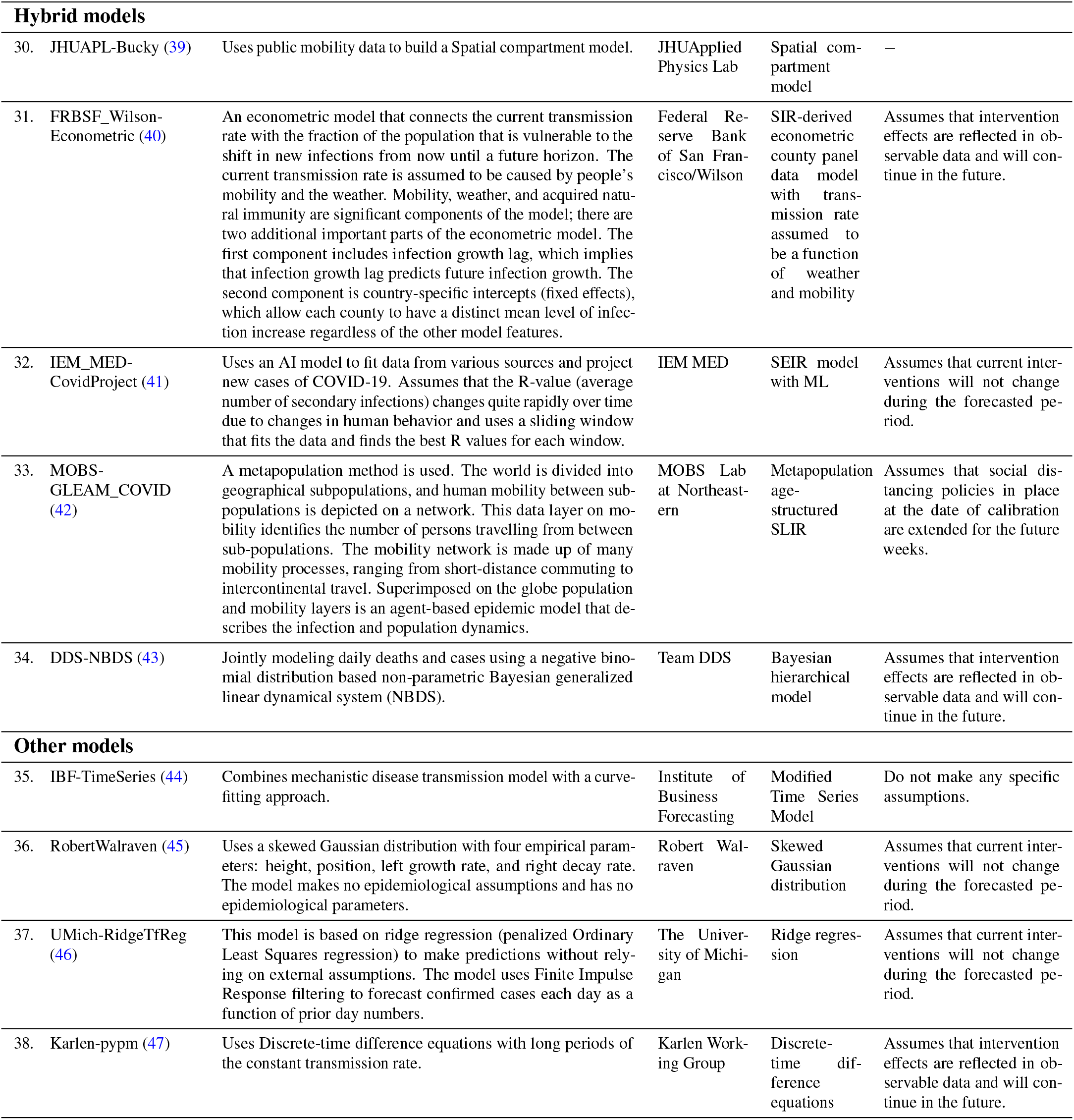
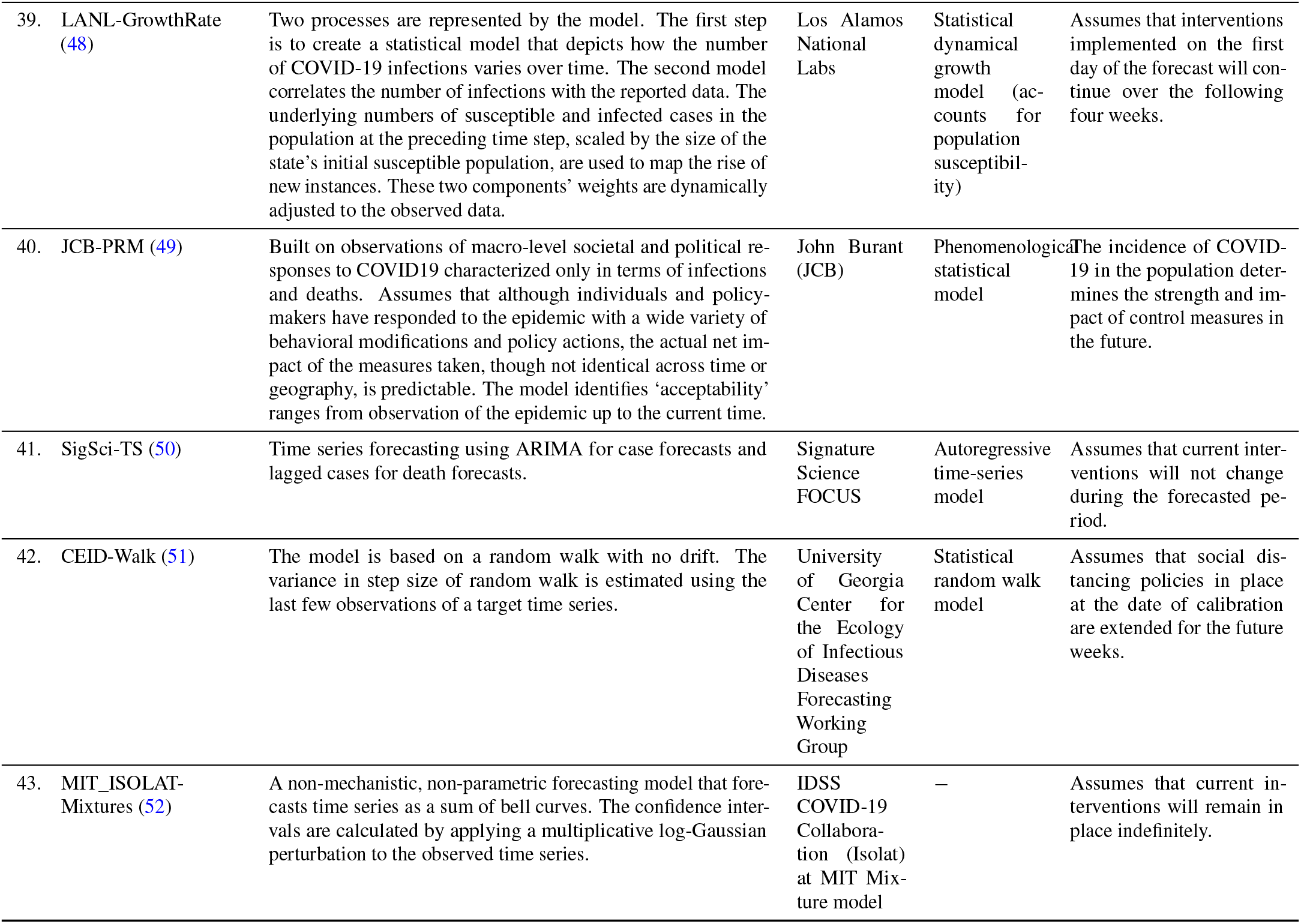
Various US-CDC COVID-19 case prediction models-their description, features employed for case prediction, the method used, assumptions made related to public health interventions.

